# Health Insurance Support on Maternal Health Care: Evidence from Survey Data in India

**DOI:** 10.1101/2020.10.20.20216093

**Authors:** Imtiyaz Ali, Saddaf Naaz Akhtar, Bal Govind Chauhan, Manzoor Ahmad Malik, Kapil Dev Singh

## Abstract

Maternal healthcare financing is key to the smooth functioning of maternal health systems in a country. In India, maternal healthcare persists as a significant public health issue. Adequate health insurance could transform the utilization of maternal health care services to prevent maternal consequences. This paper aims to examine the health insurance policies that cover maternal health and their performance in India. The unit-level social consumption data on health by the National Sample Survey Organizations (NSSO), conducted in India (2017-18), is used. Bi-variate analysis, logistic regression, and propensity scoring matching (PSM) are used to evaluate the coverage of health insurance coverage on women’s maternal health care utilization. Our findings suggest that spending on health insurance can benefit pregnant women, especially among the poor, without financial stress. The study has also minimized the financial burden and prevent high-risk pregnancy-related complications and consequences. Also, there is a need for proactive and inclusive policy development by the Government of India to promote more health insurance schemes in the public and private sectors. This can bring down the risk of maternal mortality and also boost the Indian economy in terms of a better quality of life in the long run, and the way towards more just and more egalitarian societies.

**Highlights:**

- Around 14.1% of Indian women are covered with health insurance schemes.
- Muslim women have the lowest health insurance coverage in India.
- Women covered with health insurance schemes has showed significant contributor to the better utilization of full ANC and institutional delivery compared to uncovered women in India.
- A proactive and inclusive policy development is needed by the Government of India to promote more for health insurance schemes better quality of life in the long run.

## Introduction

Health is key to economic progress and social welfare. A country with good health indicators has lower risk to fall in epidemic traps [1]. Public health challenges are thus key to address, and maternal healthcare is one such significant challenge, especially in low and middle-income countries [2,3]. But better public health measures require adequate health financing, which is very difficult to manage in developing countries. Good health spending can improve both mortality conditions and health outcomes [4]. Similarly, a robust health care system, health policies, and programs can be determinantal in addressing the health care needs of the populations.

Various studies have stressed the need for efficient health care system and the comprehensiveness of health policies to reduce health risks [5–7]. But it is difficult for low countries to manage due to limited available resources. Hence economic resources are identified to ensure the better utilization of health care without impacting much through public spending. Health insurance is one such incentive through which better healthcare utilization is achieved without impacting much on a budget of governments [5,8]. This approach can not only lower the risk of public health challenges but also reduces the public expenditure to diversify economic resources.

### Health care Financing and Maternal Health

Health spending in public health is a significant challenge for developing countries due to lack of financial resources and large health care needs [9]. Although these countries do some health spending towards their health care system, the percentage of public health expenditure is insufficient in terms of averting the health care risks they face [10]. Therefore, researchers argue for the role of financing in public health. Health financing through health insurance can be beneficial in averting maternal health care risks and key in the utilization of maternal healthcare services [11,12].

Health care financing is key to the smooth functioning of health systems in a country. Health financing through different means enables progress in health outcomes and results as adequate service coverage and financial protection [13]. Health financing has effectively done through various means ranging from revenue raising to pooling of funds and purchasing of services [14]. Since the developing countries cannot afford complete health system financing, therefore health insurance can be one key area, which can play a significant role in public-private partnerships to lower the health care risks [15]. Health financing from now on health insurance in this study is positively associated with efficient and better health care utilization. Studies in this context have found that health insurance is positively related to the use of maternal health services [16,17]. Health insurance plays a critical role in reducing the maternal mortality ratio and helps in improving maternal health outcomes [18].

Similarly, several other studies have found that the health insurance coverage increased the probability of having the recommended number of prenatal visits [19– 21]. Studies in countries like Ghana, Indonesia, and Rwanda, have found a significant positive impact of health insurance on maternal health outcomes [22,23]. Health insurance coverage lowers the risk of maternal deaths due to adequate health care financing. It provides an incentive for pregnant women to access maternal health services throughout pregnancy, childbirth, and post-delivery [24,25]. ANC services received through health insurance coverage reduces the risk for the incidence of low birth weight babies and cesarean section deliveries [26].

Similarly, it enhances the incentive for institutional births [27]. A study in Egypt while differentiating between those who access health insurance and those who do not have found that, health insurance increases maternal health care utilization. However, it has not much effect in Tanzania, where the impact has been seen very low [18].

Health insurance can not only lower the risk of maternal health but also it can be pivotal in enhancing maternal health benefits, especially in India. Studies conducted so far in India have found mixed results. A positive impact of micro health insurance on access and utilization of health services has found in Karnataka with insured individuals who were more likely to access and utilize inpatient services compared to uninsured individuals [28]. Similarly, another study has found the ineffectiveness of RSBY in reducing the burden of out-of-pocket spending on low households [29]. A study by [30] showed the most inadequate inpatient care access by low-income families even after the introduction of publicly financed insurance plans.

### Health Finance in Indian Setting

Health spending in India is a significant concern and a good reason for health distress. Public expenditure on health in India is much lower than the global average and even lower than many developing countries of Asia and Africa [31]. India’s total spending on health is only 1.28% [32]. Given the population size, this share should have been very significant. There is an urgent need for an increase in health care spending given the rising demand for health care needs, and achieving the Sustainable Development Goal (SDG) targets. The National Health Policy (2017) proposed to increase the public expenditure on health to 2.5% of GDP by 2025 and strengthen access to maternal and child health care [33,34].

Low per capita health expenditure poses a significant challenge for public health issues in India [35,36]. Looking at the current healthcare scenario, the demand for public spending on health, public health poses a considerable challenge in India’s ability to pursue better policies and address the set targets. One of the challenging aspects of managing maternal health targets, which risks the life of child and mother survival in India [35]. Even though there has been a decline in maternal and child health mortality over the past two decades, they still possess a significant threat from a public health perspective. Thus, India needs to improve its maternal health services in terms of coverage and quality to reduce maternal health risks.

Increasing health spending and better-quality care services play a crucial role in preventing life-threatening complications both during pregnancy and childbirth [5,37]. Expenditure in providing ANC, PNC, and Institutional birth services are crucial to the utilization of health care during pregnancy and childbirth. Adequate spending in delivering these services can be determinantal, and the key to reducing the maternal health risks in Indian settings [38]. Although schemes like Conditional Maternity Benefit (CMB) also called as Indira Gandhi Matritva Sahyog Yojana (IGMSY), Pradhan Mantri Surakshit Matritva Abhiyan (PMSMA), Pradhan Mantri Matru Vandana Yojana (PMMVY) and other state-level programs are launched to improve the maternal health care outcomes in India, which provide various incentives ranging from cash benefits to health care expenditure of both mother and child. But despite having these several schemes, maternal and child health care service utilization is dismal in India. Since the investment done in this context is inadequate. Thus, there has been a growing concern for executing health insurance, which can have a significant positive effect on maternal health status [22]. Adequate financing through insurance can cover the costs and also provide the means for averting early maternal health risks.

Therefore, understanding the financing, (health insurance in this paper) and its impact on maternal healthcare outcomes is central. Since any such study can be pivotal, this study will try to understand the effect of health insurance on access to and utilization of maternal healthcare services. The paper will examine the health insurance policies that cover maternal health and their performance in India since any significant finding in this context can be essential in the context of reforms and policies, that are needed to make health insurance coverage accessible to reduce maternal health risk and improve the health status of women in India.

## Materials and Methods

### Materials

The present study has used the 75^th^ round of National Sample Survey Organization (NSSO) data under the Ministry of Statistics and Program Implementation (MOSPI), conducted in India. The study has used unit data of social consumption on health, schedule 25.0 of the 75^th^ round (2017-18) by the NSSO, Government of India (July 2017 to June 2018). The 75^th^ round of NSSO data provides comprehensive information on the details of the pregnancy of every female household member aged 15-49 years who were reported to have been pregnant at any time during the last 365 days, covering prenatal care, and of childbirth and post-natal care and including expenditure on childbirth. The study has used maternal-child health care services in a 365-day reference period as a unit of analysis. A total of 113823 households (555351 individuals of which 31914 pregnant women in the last 365 days, covering prenatal care, and of childbirth and post-natal care) were successfully interviewed.

### Methods

#### Outcome variable

Three indicators of maternal health care services namely: (1) full antennal care [at least four antenatal visits, at least one tetanus toxoid (TT) injection and iron-folic acid tablets or syrup was taken for 100 or more days], (2) institutional delivery [delivery conducted in the health facility irrespective of the type of institution], and (3) postnatal care [women received check-ups in the first 7 days after the delivery of the child] was the outcome variable of the study. All of these variables are made dichotomous (coded as 0 or 1).

#### Predictor variables

The main independent variable of interest has been health insurance coverage. The NSSO asked respondents whether they were covered by health insurance for health insurance support and what type of health insurance they had. We constructed a dichotomous variable of whether a woman was covered by any health insurance. Also, this has been taken as treatment variable in the PSM analysis. Additionally, age of the respondent (<20 years, 20-24 years, 25-29 years, ≥ 30 years), place of residence (Rural, Urban), religion (Hindu, Muslim, Christianity, Others), Social Group (STs, SCs, OBC, Others), household size (≤ 5 members, > 5 members), wealth quintile (Poorest, Poor, Middle, Rich, Richest) mother’s education (Illiterate, Primary, Secondary, Higher), mother’s working status (Working, Not working) have been used as a predictor variable in the study.

#### Statistical analysis

The bivariate and multivariate statistical techniques were used to analyze the data. In the multivariate technique, logistic regression and propensity scoring matching (PSM) was used to evaluate the effect or consequences of the coverage of health insurance on women’s maternal health care utilization.

##### Logistic regression

Logistic regression was carried out to see the significant effect of predictor variables on outcome variable. The general regression model used for the study has been defined in equation below:

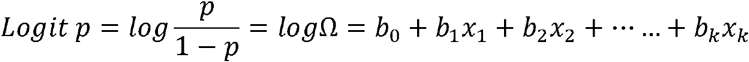

Where, where *p* is the probability that the event Y occurs, coefficient ‘*b*’ is the factor by which the odds changes with unit increase in independent variable. If “*b*” is positive, odds ratio will increase, as this factor will be greater than 1. Contrary to that if ‘*b*’ is negative, odd ratio will decrease. When ‘*b*’ is 0, the factor exponential of ‘*b*’ equal to 1 and, therefore the odds remain unchanged.

##### Propensity score matching (PSM)

Our study adopted PSM analysis to estimate the effect of the health insurance coverage on maternal healthcare utilization - i.e., full ANC, Institutional delivery and post-natal care. PSM has been a useful tool that can evaluate the treatment effect for cross-sectional/observational/non-experimental data [39].

In the present study, women who seek maternal health care utilization may be more likely to enrol in health insurance (endogeneity). Also, the sample of women who were enrolled in health insurance was not a random selection of women. The positive effects of health insurance may be overstated using ordinary regression even if these factors are controlled for in the model because selection bias can result when the distribution of the characteristics of women with health insurance and those without health insurance differ [40]. Besides, women who are enrolled in health insurance and those who do not may vary in their aversion to risk. Risk-averse women are both more likely to seek health insurance coverage and seek maternal health care services utilization. This unobserved heterogeneity in women’s characteristics results with unobserved self-selection bias. Therefore, the strength of PSM is that it is not affected by selection bias like a typical regression model.

For the present study, we used a binary exposure variable (women covered by health insurance: Yes, vs No), and all possible available confounding covariates (independent treatment variables) that are determinants of both exposure and response were adjusted.

With this approach, we have calculated the robust estimators to determine the effect of health insurance on maternal health care services (full ANC, institutional delivery and PNC). The main assumption in this method is that conditionality of propensity score; the observable selected characteristics of the treatment (insured) and control groups (uninsured) have similar distributions [41]. This assumption t-test was applied by using ‘pscore’ command [42]. After satisfying the balancing property, we estimated the ‘teffect’ to obtain the average exposure effect (AEE) on insured uninsured women at individual levels. In this paper, we present results using the nearest neighbourhood propensity score method; in the matching, estimator sorts all records by the estimated propensity score and then searches forward and backward for the closet control groups. The study has used STATA 14.0 package for the complete analysis.

## Results

Fig.1 presents the percentage of women covered by a specific type of health insurance support in India. Women who have been covered health insurance support has found to be 14.1% at the national level during 2017-18 (Fig.1). There has been a specific type of health insurance supports in India in the period 2017-18. Around 85.9% of women had not undergone any health insurance support in India. The Fig.1 also depicts that the highest health insurance supports have been observed with around 10.1% among women who opted the Government-sponsored support (such as RSBY, Arogyasari, etc.). While the health insurance supports arranged by households with insurance companies have been found to be 0.8% only. In the case of Government/PSU employer, it has been found to be only 1% coverages whereas Employer supported showed 1.1%.

**Fig. 1.**
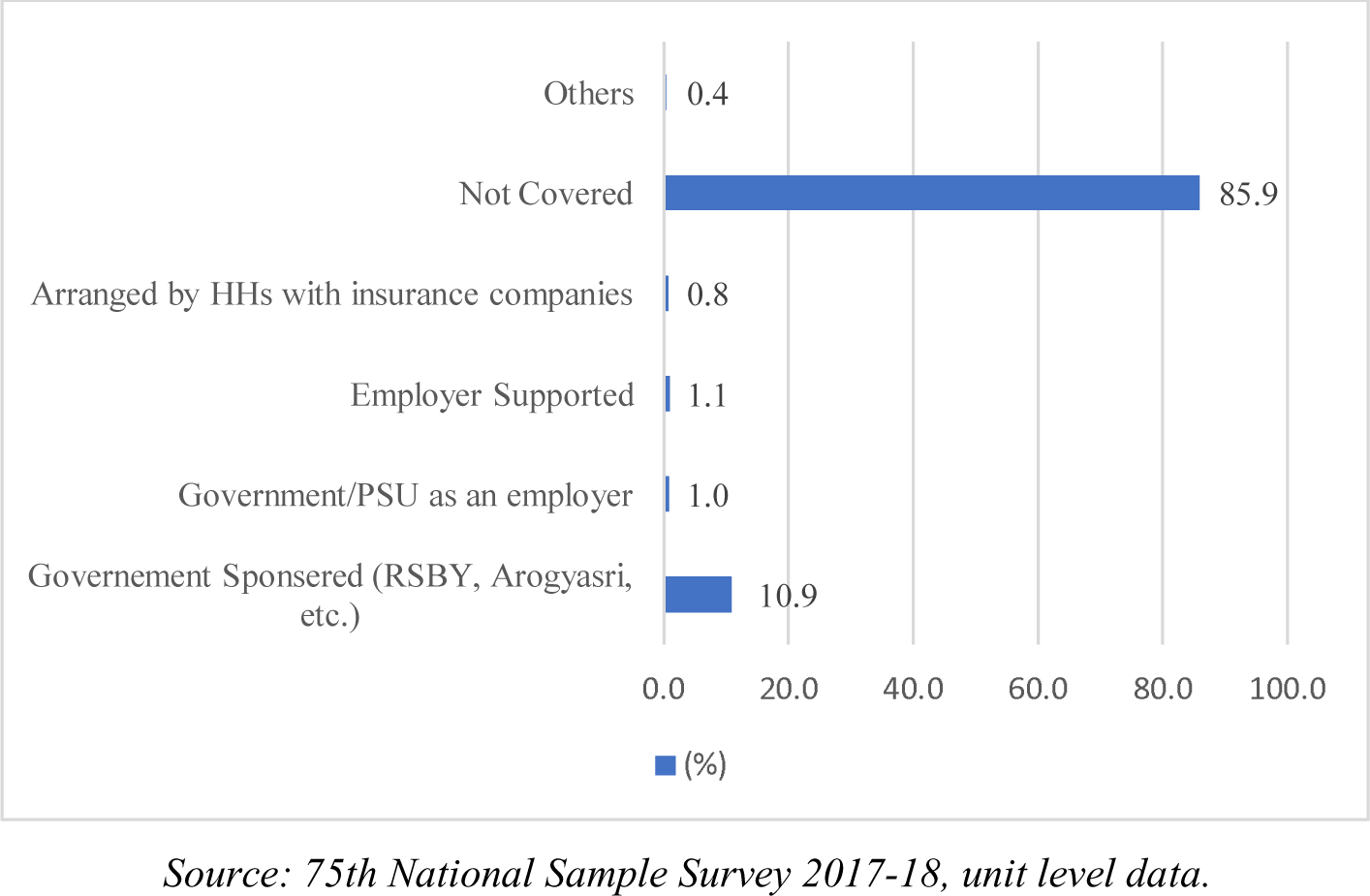
Percentage of women covered by specific type of health insurance supports in India.

At the national level, the rural insured women are found to 10% lower than urban insured women in receiving full ANC and similarly is observed in case of uninsured women. High percentage of institutional delivery is observed in which around 95% insured women reside in rural areas and 99% reside in urban. While receiving utilization of PNC, the percentage of insured and uninsured women are found to be same in rural area (approx 88%) and lesser than the urban insured and uninsured women in India.

Table 1 showed the percentage of women who have received maternal healthcare services in India in the period 2017-18. The results in the Table 1 shows sample distribution, insurance coverage, and utilization of maternal healthcare services of 31914 pregnant women (any time in the last 365 days) by selected socioeconomic characteristics. Results from the table shows that around 14.1% women are covered with health insurance, and 85.9% of women are uncovered. Health insurance coverage is found to be greater in urban areas (16.6%) than in rural areas (13.3%). Similarly, health insurance coverage of women along with other socio-economic and demographic variables can be found in table 1. Table 1 also shows the selected maternal healthcare services (full ANC, institutional delivery, and PNC) along with various socio-economic and demographic characteristics.

**Table 1.** Percentage of women received the selected utilization of maternal health services in India, 2017-18.

| Background | Sample distribution |  | Insurance coverage | Utilization of maternal health services |  |  |
| --- | --- | --- | --- | --- | --- | --- |
|  | % | n |  | Full ANC (%) | Institutional delivery (%) | PNC (%) |
| *Place of Residence* |  |  |  |  |  |  |
| Rural | 75.9 | 19104 | 13.3 | 32.6 | 91.9 | 88.0 |
| Urban | 24.1 | 12810 | 16.6 | 45.0 | 96.3 | 92.7 |
| *Religion* |  |  |  |  |  |  |
| Hindu | 79.0 | 23869 | 14.7 | 35.6 | 93.8 | 89.3 |
| Muslim | 16.8 | 4700 | 8.47 | 33.3 | 88.9 | 87.2 |
| Christian | 2.2 | 2113 | 37.5 | 49.4 | 90.5 | 90.9 |
| Others | 2.0 | 1232 | 8.6 | 41.5 | 94.9 | 94.9 |
| **Insurance Status** |  |  |  |  |  |  |
| Covered | 14.1 | 5547 | 14.1 | 47.0 | 96.1 | 89.9 |
| Uncovered | 85.9 | 26367 | 0.0 | 33.8 | 92.4 | 89.0 |
| **Social Group** |  |  |  |  |  |  |
| ST | 10.1 | 4693 | 23.4 | 35.3 | 89.3 | 88.9 |
| SC | 21.0 | 5657 | 10.9 | 33.7 | 92.7 | 89.3 |
| OBC | 46.1 | 12831 | 14.0 | 34.8 | 92.5 | 88.3 |
| Others (or General) | 22.9 | 8733 | 13.1 | 42.8 | 95.7 | 90.6 |
| **Household Size** |  |  |  |  |  |  |
| ≤5 members | 52.9 | 17479 | 15.6 | 36.5 | 92.8 | 89.5 |
| > 5 members | 47.1 | 14435 | 12.4 | 34.6 | 93.0 | 88.7 |
| **Wealth Quintile** |  |  |  |  |  |  |
| Poorest | 30.8 | 7212 | 8.4 | 24.8 | 89.7 | 86.5 |
| Poor | 24.4 | 6555 | 10.6 | 32.5 | 92.5 | 87.6 |
| Middle | 20.3 | 6491 | 18.3 | 41.4 | 93.4 | 89.9 |
| Rich | 14.8 | 6180 | 18.3 | 45.9 | 96.8 | 92.0 |
| Richest | 9.8 | 5476 | 25.4 | 50.4 | 98.1 | 95.6 |
| **Woman's Education** |  |  |  |  |  |  |
| Illiterate | 19.7 | 4090 | 11.7 | 23.4 | 84.1 | 83.2 |
| Primary | 39.0 | 11640 | 12.5 | 35.5 | 94.0 | 89.0 |
| Secondary | 28.6 | 10350 | 15.1 | 39.2 | 96.0 | 91.9 |
| Higher | 12.8 | 5834 | 20.3 | 46.8 | 97.6 | 93.1 |
| **Women's Working Status** |  |  |  |  |  |  |
| Working | 10.1 | 3218 | 23.5 | 34.9 | 91.6 | 84.4 |
| Not Working | 89.9 | 28696 | 13.0 | 35.7 | 93.1 | 89.6 |
| **Woman's Age** |  |  |  |  |  |  |
| Less than 20 years | 4.5 | 956 | 16.6 | 33.5 | 92.7 | 90.7 |
| 20-24 years | 39.7 | 10634 | 13.9 | 35.1 | 94.0 | 89.1 |
| 25-29 years | 35.5 | 12137 | 14.6 | 38.2 | 93.2 | 88.9 |
| 30 and above | 20.3 | 8187 | 13.1 | 32.8 | 90.7 | 89.2 |
| Total | 100 | 31914 | 14.1 | 35.6 | 93.0 | 89.1 |

Notes: ST: Schedule Tribe; SC: Schedule Caste; OBC; Other backward caste.
Source: 75<sup>th</sup> Round National Sample Survey 2017-18, unit level data.

Table 2 shows the percentage of women covered and uncovered by health insurance support who received the selected maternal healthcare services (full ANC, institutional delivery, and PNC) in India, 2017-18. The results showed that greater percentage of women in urban areas are covered by health insurance support. Similarly, it can be seen that household size plays a key role in health insurance coverage, with women having less than 5 members have higher coverage of health insurance. Similarly factors like income, education and early age are key for utilising the maternal health care services though health insurance as shown in the table 2.

**Table 2.** Percentage of women covered and uncovered by health insurance supports who received the all selected utilization of maternal health services in India, 2017-18.

| Background | Full ANC |  | Inst. Delivery |  | PNC |  |
| --- | --- | --- | --- | --- | --- | --- |
|  | Covered by health insurance support | Uncovered by health insurance support | Covered by health insurance support | Uncovered by health insurance support | Covered by health insurance support | Uncovered by health insurance support |
| **Place of Residence** |  |  |  |  |  |  |
| Rural | 44.2 | 30.7 | 94.8 | 91.5 | 87.9 | 88.0 |
| Urban | 54.0 | 43.3 | 99.1 | 95.8 | 94.5 | 92.3 |
| **Religion** |  |  |  |  |  |  |
| Hindu | 47.4 | 33.5 | 95.8 | 93.5 | 90.2 | 89.2 |
| Muslim | 38.8 | 32.9 | 98.4 | 88.1 | 85.4 | 87.3 |
| Christian | 54.6 | 46.2 | 96.6 | 86.8 | 93.0 | 89.8 |
| Others | 51.5 | 40.6 | 96.2 | 94.7 | 93.0 | 95.0 |
| **Caste** |  |  |  |  |  |  |
| ST | 40.2 | 32.5 | 91.2 | 88.6 | 85.6 | 89.6 |
| SC | 51.0 | 30.8 | 96.4 | 92.2 | 90.4 | 89.1 |
| OBC | 46.1 | 32.1 | 97.8 | 91.7 | 90.0 | 88.1 |
| Others (or General) | 51.3 | 40.3 | 96.7 | 95.5 | 92.9 | 90.3 |
| **Household size** |  |  |  |  |  |  |
| ≤ 5 members | 47.8 | 34.4 | 96.0 | 92.3 | 94.0 | 88.9 |
| > 5 members | 45.8 | 33.0 | 96.4 | 92.6 | 84.6 | 89.2 |
| **Wealth quintile** |  |  |  |  |  |  |
| Poorest | 29.2 | 24.4 | 94.4 | 89.3 | 85.2 | 86.6 |
| Poor | 44.2 | 31.1 | 96.5 | 92.0 | 83.5 | 88.1 |
| Middle | 46.9 | 40.2 | 95.1 | 93.1 | 90.4 | 89.8 |
| Rich | 56.1 | 43.6 | 96.2 | 97.0 | 94.3 | 91.5 |
| Richest | 58.5 | 46.7 | 99.0 | 97.8 | 96.4 | 95.3 |
| **Women's education** |  |  |  |  |  |  |
| Illiterate | 25.8 | 23.1 | 93.6 | 82.9 | 84.1 | 83.0 |
| Primary | 46.8 | 33.9 | 94.5 | 93.9 | 88.9 | 89.1 |
| Secondary | 52.9 | 36.8 | 97.8 | 95.7 | 91.6 | 91.9 |
| Higher | 56.4 | 44.3 | 98.8 | 97.3 | 95.0 | 92.6 |
| **Women's working Status** |  |  |  |  |  |  |
| Working | 45.2 | 31.7 | 91.8 | 91.5 | 85.1 | 84.1 |
| Not Working | 47.4 | 34.0 | 97.1 | 92.6 | 90.9 | 89.4 |
| **Mother's age** |  |  |  |  |  |  |
| Less than 20 years | 47.0 | 30.8 | 96.1 | 92.1 | 91.2 | 90.8 |
| 20-24 years | 44.9 | 33.5 | 96.6 | 93.6 | 88.9 | 89.1 |
| 25-29 years | 49.2 | 36.3 | 95.7 | 92.9 | 90.7 | 88.6 |
| 30 and above | 47.1 | 30.6 | 96.1 | 90.0 | 90.2 | 89.1 |
| Total | 47.0 | 33.8 | 96.1 | 92.5 | 89.9 | 89.0 |

Notes: ST: Schedule Tribe; SC: Schedule Caste; OBC; Other backward caste.
Source: 75<sup>th</sup> Round National Sample Survey 2017-18, unit level data.

Table 3 presented the results of logistic regression showing the odds ratio and 95% confidence interval (CI) of health insurance status on all selected maternal healthcare services in India in the period 2017-18 by selected socioeconomic characteristics. About 11% of urban women are less likely to have health insurance supports in terms of full ANC services compared to rural women while on the other side, urban women are 38% more likely to have institutional delivery compared to rural women. The odds of receiving full ANC services and institutional delivery are less significant (OR=0.92, [0.86-0.99] and (OR=0.50, [0.41-0.56]) among Muslim women compared to Hindu women. The odds of Christian women are about 32% (full ANC), 65% (institutional delivery), and 45% (PNC) lesser significant than the odds of Hindu women. The uncovered insured women are found to be 33% lesser significant who are receiving full ANC and 56% lesser significant who are receiving institutional delivery compared to covered insured women. The results show that socio-economic characteristics play a significant role in utilizing health care benefits through health insurance support. Since the odds ratio for women with better socio-economic conditions like income, education, caste religion and so on were higher than those belong to vulnerable groups when analyzing the insurance support as shown by the results in the table 3.

**Table 3.** Result of logistic regression showing the odds of the selected utilization of maternal health services in India, 2017-18.

| Background | Full ANC | Institutional<br>Delivery | PNC |
| --- | --- | --- | --- |
| **Place of Residence** |  |  |  |
| Rural® | 1.00 | 1.00 | 1.00 |
| Urban | 0.89† [0.84-0.93] | 1.38† [1.16-1.58] | 0.98 [0.89-1.07] |
| **Religion** |  |  |  |
| Hindu | 1.00 | 1.00 | 1.00 |
| Muslim | 0.92†† [0.86-0.99] | 0.50† [0.41-0.56] | 1.08 [0.95-1.22] |
| Christian | 0.68† [0.62-0.77] | 0.36† [0.35-0.53] | 0.55† [0.47-0.65] |
| Others | 0.77† [0.69-0.88] | 0.37† [0.29-0.47] | 0.67† [0.55-0.81] |
| **Insurance Status** |  |  |  |
| Covered® | 1.00 | 1.00 | 1.00 |
| Uncovered | 0.67† [0.63-0.71] | 0.44† [0.37-0.55] | 0.93 [0.83-1.04] |
| **Social Group** |  |  |  |
| ST | 1.00 | 1.00 | 1.00 |
| SC | 1.37† [1.26-1.50] | 1.88† [1.62-2.38] | 1.14 [0.98-1.31] |
| OBC | 1.25† [1.15-1.36] | 1.93† [1.81-2.55] | 1.05 [0.92-1.20] |
| Others (or General) | 1.30† [1.20-1.43] | 2.18† [1.91-2.85] | 1.00 [0.86-1.15] |
| **Household Size** |  |  |  |
| ≤5 members® | 1.00 | 1.00 | 1.00 |
| > 5 members | 1.04 [0.99-1.09] | 1.23†† [1.09-1.39] | 0.86† [0.80-0.94] |
| **Wealth Quintile** |  |  |  |
| Poorest | 1.00 | 1.00 | 1.00 |
| Poor | 1.37† [1.28-1.48] | 1.44† [1.15-1.54] | 1.02 [0.92-1.15] |
| Middle | 1.83† [1.70-1.97] | 1.64† [1.34-1.88] | 0.99 [0.88-1.11] |
| Rich | 2.28† [2.12-2.47] | 2.52† [1.93-2.96] | 1.37† [1.20-1.57] |
| Richest | 2.38† [2.18-2.61] | 3.03† [2.15-3.83] | 1.87† [1.57-2.23] |
| **Woman's's Education** |  |  |  |
| Illiterate® | 1.00 | 1.00 | 1.00 |
| Primary | 1.58† [1.46-1.72] | 2.15† [1.93-2.54] | 1.49† [1.33-1.67] |
| Secondary | 1.81† [1.67-1.97] | 3.34† [3.00-4.25] | 2.04† [1.80-2.31] |
| Higher | 2.19† [2.00-2.41] | 5.09† [4.18-7.42] | 2.41† [2.05-2.83] |
| **Woman's Working Status** |  |  |  |
| Working | 1.00 | 1.00 | 1.00 |
| Not Working | 1.05 [0.97-1.14] | 1.61† [1.39-1.91] | 1.10 [0.97-1.25] |
| **Woman's Age** |  |  |  |
| Less than 20 years | 1.00 | 1.00 | 1.00 |
| 20-24 years | 0.96 [0.84-1.1] | 0.94 [0.66-1.33] | 0.82 [0.64-1.06] |
| 25-29 years | 0.91 [0.79-1.04] | 0.85 [0.6-1.21] | 0.88 [0.68-1.13] |
| 30 and above | 0.85†† [0.74-0.98] | 0.81 [0.57-1.16] | 1.03 [0.80-1.34] |

Notes: Significance level-†p<0.01, ††p<0.05, €p<0.1, ®Reference category; ST: Schedule Tribe; SC: Schedule Caste; OBC; Other backward caste. Source: 75<sup>th</sup> Round National Sample Survey 2017-18, unit level data.

Propensity Score Matching (PSM) technique was used for furthering the analysis to understand the effect of health insurance support/health insurance on maternal health care service utilization. Table 4 shows the basic descriptive statistics of the estimated propensity score matching results all the cases. The results in the table shows the balancing property at 1% significance level. The score indicated that there were no systematic differences in covariates between women at individual level, covered and uncovered by health insurance status/health insurance support. This means that if both group of women had similar socio-demographic characteristics except for health insurance support, then a difference in means in the utilization of maternal health care services between women of exposed and of non-exposed groups could be attributed to health insurance.

**Table 4.**
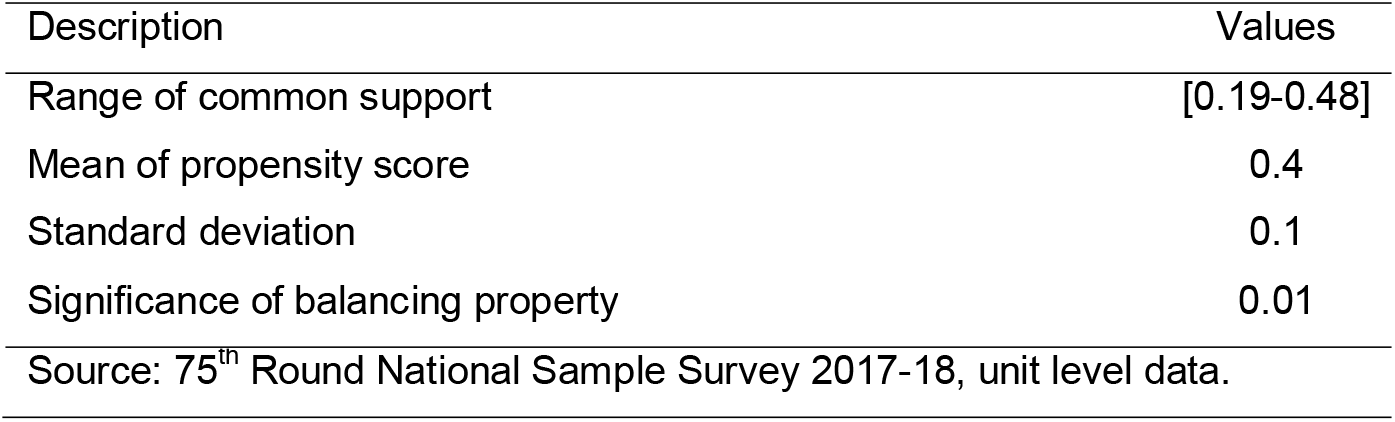
Description of propensity score.

Table 5 showed the AEE for ANC, institutional delivery and PNC with the nearest neighbour matching technique. The study showed that there had been a significant positive exposure effect of women covered with health insurance supports of 7.2% to receive full ANC, 9.1% to receive institutional delivery, and 2.8% to receive PNC in India. The results in Table 5, reflected the hypothesis that health insurance support plays a significant role in accessing and utilizing the better maternal health care services which can lower the risk for maternal and child health burden in India.

**Table 5.**
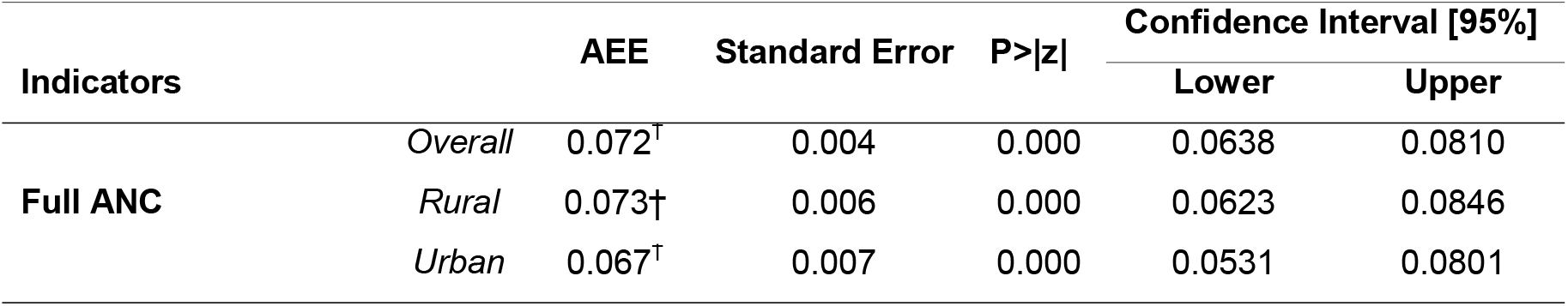
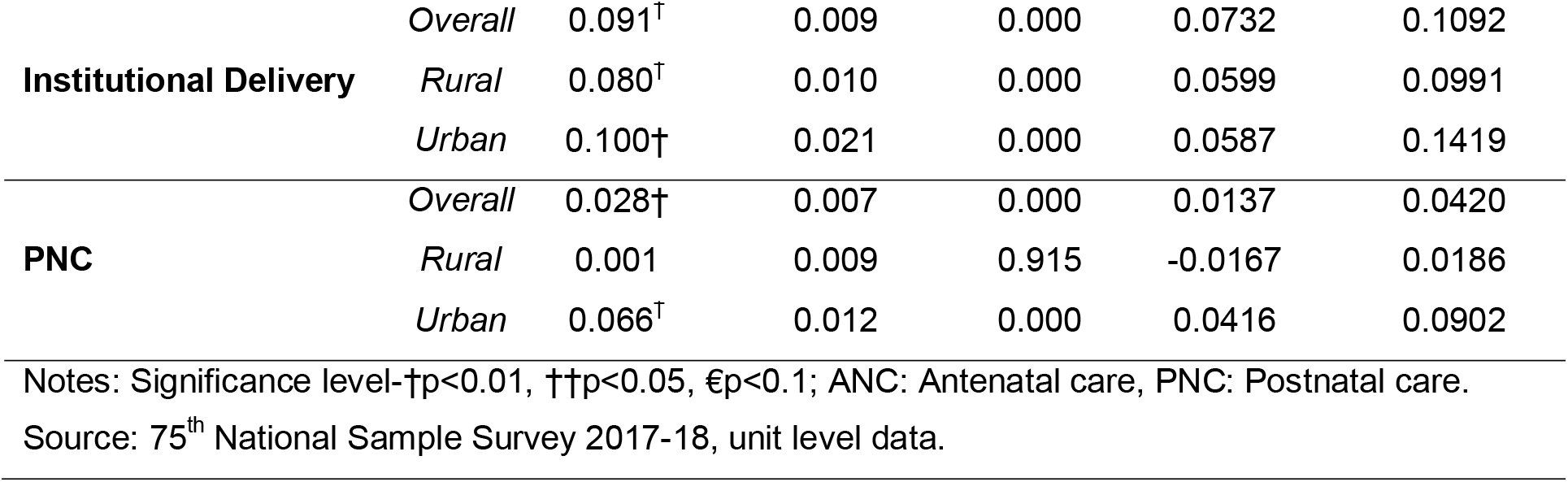
Average exposure effect (AEE) for MCH services of women covered with and without health insurance supports in India by place of residence, 2017-18.

| Indicators |  | AEE | Standard Error | P>\|z\| | Confidence Interval [95%] |  |
| --- | --- | --- | --- | --- | --- | --- |
|  |  |  |  |  | Lower | Upper |
| Full ANC | Overall | 0.072<sup>†</sup> | 0.004 | 0.000 | 0.0638 | 0.0810 |
|  | Rural | 0.073† | 0.006 | 0.000 | 0.0623 | 0.0846 |
|  | Urban | 0.067<sup>†</sup> | 0.007 | 0.000 | 0.0531 | 0.0801 |
| **Institutional Delivery** | *Overall* | 0.091<sup>†</sup> | 0.009 | 0.000 | 0.0732 | 0.1092 |
|  | *Rural* | 0.080<sup>†</sup> | 0.010 | 0.000 | 0.0599 | 0.0991 |
|  | *Urban* | 0.100<sup>†</sup> | 0.021 | 0.000 | 0.0587 | 0.1419 |
| **PNC** | *Overall* | 0.028<sup>†</sup> | 0.007 | 0.000 | 0.0137 | 0.0420 |
|  | *Rural* | 0.001 | 0.009 | 0.915 | -0.0167 | 0.0186 |
|  | *Urban* | 0.066<sup>†</sup> | 0.012 | 0.000 | 0.0416 | 0.0902 |

Notes: Significance level-†p<0.01, ††p<0.05, €p<0.1; ANC: Antenatal care, PNC: Postnatal care.
Source: 75<sup>th</sup> National Sample Survey 2017-18, unit level data.
Source: 75th National Sample Survey 2017-18, unit level data.

## Discussion

Assessing maternal health outcomes has been an essential aspect of the public health domain. Multiple studies have examined the range of issues related to maternal health, varying from access, utilization, inequality, and benefits [43]. But the role of health financing has been a challenging one to examine. It is a well-accepted fact now that health health insurance plays a significant role in improving the financial protection and utilization of maternal healthcare system [16–18,22– 24,26,44–46]. But studying it in maternal health outcomes is challenging. Since a pregnant woman needs continuous medical attention from the moment they conceive and involves cost from the early gestation periods [47], which is not viable for everyone especially the women from developing countries context [48–50]. While evaluating the impact of the health insurance status on maternal health care utilization in India, we found some significant and positive association of health care financing through health insurance in terms of maternal health outcomes. We found that with improvement in maternal health indicators like Institutional delivery, ANC, and PNC utilization over the past decade, India is significantly benefiting from the health care financing in terms of maternal health care utilization. Institutional delivery was now increased to more than 90% from 83% in 2014, which leads to rising ANC and PNC. Hence, this reflects the fact that public financing has significantly contributed to the better and increased utilization of health care services [51]. While examining the role of health insurance in utilizing maternal health care benefits, we found that women covered through insurance benefits are more likely to access these benefits as compared to uninsured women. Our findings were consistent with many earlier works in this aspect [16,17,22,23,46].

Our study signifies the role of public financing, as the results indicate that the increase in health financing through health insurance has positive consequences, but reflects the need for enhancing public health financing. Since funding of health care costs and support during and preceding the weeks after childbirth of women has a positive impact on maternal health outcomes [52]. The study shows that health financing has increased in India over time, but it has not been significant, mainly due to knowledge gaps and inadequate policy responses. There is a lack of a target-oriented approach in the current policies, which has resulted in these policy gaps and the vulnerability of low socio-economic groups in India. However, the coverage has been better among the households with better socio-economic conditions as reflected by earlier studies in this context [53,54].

Health insurance coverage is vital in reducing the health disparities since it increases the incentive for utilizing the health care benefits of women and also reduce the health risk [55]. Our results indicate the advantage of better socio-economic groups, particularly those benefitting from the health insurance coverage. It has now understood that women’s better income and educational households benefit from health insurance, as observed in the PSM results. Huge variation was observed among women covered and uncovered by health insurance in maternal health care utilization by socio-economic and demographic characteristics as followed by similar studies earlier [21,23,44,56–59]. To sum up, it is evident from our study that health insurance has a significant role to play not only in the maximization of maternal health care utilization but also through the betterment of maternal health care services. Therefore, government policies aiming at incentivizing public financing through health insurance can shrink the challenges of public health burden and minimize the risks while averting the maternal and child health losses.

## Conclusions

Coverage of health insurance is the most significant contributor to the better utilization of full ANC and institutional delivery at the national level. It can not only maximize the utilization but also increase the overall welfare by reducing public health burden. Health insurance through public spending can play a significant role in achieving health targets and SDG goals since they are significantly associated with better utilization of health care services. Our findings suggest that spending on health insurance can benefit pregnant women, especially among poor without any financial stress, and this would also minimize the financial burden and prevent high-risk pregnancy-related complications and consequences. Also, there is a need for proactive and inclusive policy development by the Government of India to promote more for health insurance schemes in both the public and private sectors. This can bring down the risk of maternal mortality and also boost the Indian economy in terms of a better quality of life in the long run, and the way towards more just and more egalitarian societies.

## Data Availability

The present study has used the 75th round of National Sample Survey Organization (NSSO) data under the Ministry of Statistics and Program Implementation (MOSPI), conducted in India. The study has used unit data of social consumption on health, schedule 25.0 of the 75th round (2017-18) by the NSSO, Government of India (July 2017 to June 2018).
This data is openly available for the study.

http://www.mospi.gov.in/unit-level-data-report-nss-75th-round-july-2017-june-2018-schedule-250social-consumption-health

## Appendix

**Fig. 2.**
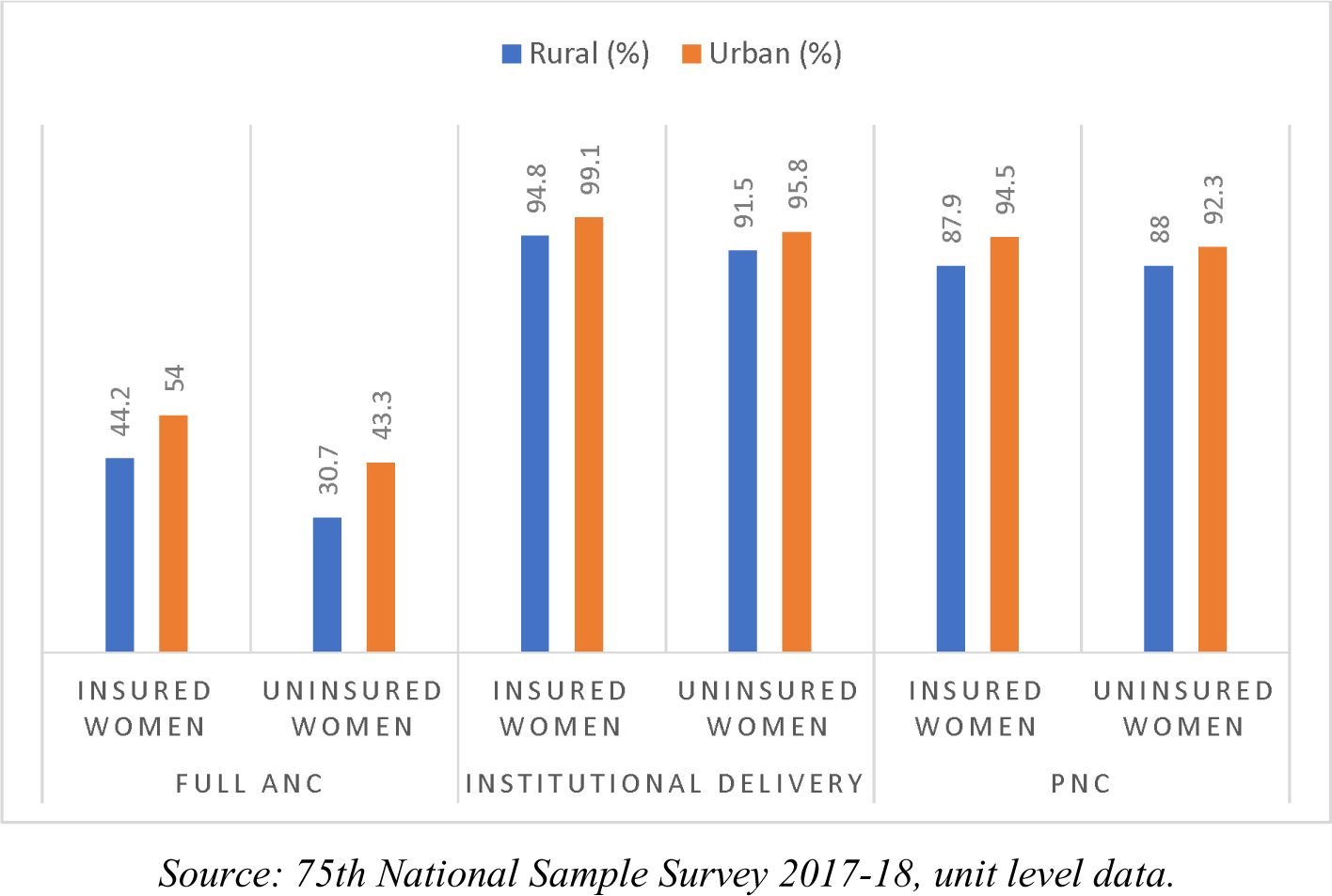
Percentage of women covered and uncovered by health insurance supports who received the all selected utilization of maternal health services by place of residence in India, 2017-18.

